# Fine analysis of lymphocyte subpopulations in SARS-CoV-2 infected patients: toward a differential profiling of patients with severe outcome

**DOI:** 10.1101/2021.08.31.21262538

**Authors:** Giovanna Clavarino, Corentin Leroy, Olivier Epaulard, Tatiana Raskovalova, Antoine Vilotitch, Martine Pernollet, Chantal Dumestre-Pérard, Federica Defendi, Marion Le Marechal, Audrey Le Gouellec, Pierre Audoin, Jean-Luc Bosson, Pascal Poignard, Matthieu Roustit, Marie-Christine Jacob, Jean-Yves Cesbron

**Affiliations:** Laboratoire d’Immunologie, Pôle de Biologie, Centre Hospitalier Universitaire Grenoble Alpes, Grenoble, France; Cellule d’Ingénierie des Données, Centre Hospitalier Universitaire Grenoble Alpes, Grenoble, France; Centre d’Investigation Clinique de l’Innovation et de la Technologie (CIC-IT), Centre Hospitalier Universitaire Grenoble Alpes, Grenoble, France; Service de Maladies Infectieuses, Centre Hospitalier Universitaire Grenoble Alpes, Grenoble, France; Université Grenoble Alpes, CNRS, Grenoble INP, TIMC, Grenoble, France; Université Grenoble Alpes, CNRS, CEA, IBS, Grenoble, France; Service de Biochimie Biologie Moléculaire et Toxicologie Environnementale, Pôle de Biologie, Centre Hospitalier Universitaire Grenoble Alpes, Grenoble, France; Unité recherche, Pôle de Biologie, Centre Hospitalier Universitaire Grenoble Alpes, Grenoble, France; Laboratoire de Virologie, Pôle de Biologie, Centre Hospitalier Universitaire Grenoble Alpes, Grenoble, France; Centre d’Investigation Clinique INSERM CIC1406, Centre Hospitalier Universitaire Grenoble Alpes, Grenoble, France; Université Grenoble Alpes, INSERM, UMR 1300, HP2, Grenoble, France

## Abstract

COVID-19 is caused by the human pathogen severe acute respiratory syndrome coronavirus 2 (SARS-CoV-2) and has resulted in widespread morbidity and mortality. CD4^+^ T cells, CD8^+^ T cells and neutralizing antibodies all contribute to control SARS-CoV-2 infection. However, heterogeneity is a major factor in disease severity and in immune innate and adaptive responses to SARS-CoV-2. We performed a deep analysis by flow cytometry of lymphocyte populations of 125 hospitalized SARS-CoV-2 infected patients on the day of hospital admission. Five clusters of patients were identified using hierarchical classification on the basis of their immunophenotypic profile, with different mortality outcomes. Some characteristics were observed in all the clusters of patients, such as lymphopenia and an elevated level of effector CD8^+^CCR7^-^ T cells. However, low levels of T cell activation are associated to a better disease outcome; on the other hand, profound CD8^+^ T-cell lymphopenia, a high level of CD4^+^ and CD8^+^ T-cell activation and a high level of CD8^+^ T-cell senescence are associated with a higher mortality outcome. Furthermore, a cluster of patient was characterized by high B-cell responses with an extremely high level of plasmablasts. Our study points out the prognostic value of lymphocyte parameters such as T-cell activation and senescence and strengthen the interest in treating the patients early in course of the disease with targeted immunomodulatory therapies based on the type of adaptive response of each patient.

## Introduction

Coronavirus disease 2019 (COVID-19) is caused by the human pathogen severe acute respiratory syndrome coronavirus 2 (SARS-CoV-2) and has resulted in widespread morbidity and mortality. The total number of lymphocytes, CD4^+^ T cells, CD8^+^ T cells and natural killer cells significantly decreases in COVID-19 patients, with the lowest levels in severe cases [1]; in particular, a decrease in CD8^+^ T cells and an increase in plasmablasts in infected patients have been described [2]. COVID-19 severity and duration seem to be dependent on the early evasion of innate immune recognition, especially with defects in type 1 interferon pathways [2], and the subsequent kinetics of the adaptive immune response [3]. Since severe COVID-19 is associated with high levels of IL-6, sepsis has been used as a prototype of critical illness for the understanding of severe COVID-19 pathogenesis. However, even if a low expression of human leucocyte antigen D related (HLA-DR) on CD14^+^ monocytes has been described in some patients, this pattern is distinct from the immunoparalysis state reported in bacterial sepsis or severe respiratory failure caused by influenza [4] [5].

CD4^+^ T cells, CD8^+^ T cells and neutralizing antibodies all contribute to control SARS-CoV-2 infection. However, heterogeneity is a major factor in disease severity and in immune innate and adaptive responses to SARS-CoV-2. Deep immune profiling of lymphocyte populations has been performed by using high dimensional flow cytometry, leading to the definition of different immunotypes: immunotype 1 characterized by CD4^+^ T cell activation, exhausted CD8^+^ T cells, presence of plasmablasts and associated with more severe disease, immunotype 2 characterized by less CD4^+^ T cell activation, the presence of effector CD8^+^ subsets and proliferating memory B cells, and immunotype 3 with minimal lymphocyte activation response and negatively associated with disease severity [6] [7]. In another study conducted with principal components analysis and hierarchical clustering, a vast array of immunological parameters has been measured, with the description of three distinct phenotypes: a humoral response deficiency phenotype, a hyper-inflammatory phenotype and a complement-dependent phenotype [8]. In the present study, we performed a fine analysis of lymphocyte subsets of SARS-CoV-2 infected hospitalized patients on the day of admission in order to better characterize the adaptive immune response and possibly define patient trajectories with different disease progression courses.

## Methods

### Ethics statements

Overall 146 patients infected with SARS-CoV-2 were recruited in Grenoble Alpes University Hospital between March and September 2020, either in a retrospective (n=127) or in a prospective (n=19) study. Clinical and biological data were fully available for 125 patients (S1 Fig). The study was performed in accordance with the Declaration of Helsinki, Good Clinical Practice guidelines and CNIL (Commission Nationale de l’Informatique et des Libertés) methodology reference. Patients were informed and non-opposition (BioMarCoViD retrospective study) or written consent (AcNT-COVID-19 prospective study) was obtained, according to French law. Ethical approval for the prospective study was given by the relevant ethics committee (Comité de Protection des Personnes Ile de France II, N°IDRCB: 2020-A00904-35) and registered in clinicaltrial.gov (NCT04596098).

Laboratory confirmation for SARS-CoV-2 was defined as a positive result of real-time reverse transcriptase-polymerase chain reaction assay of nasopharyngeal swabs. Follow-up of the patients at days 3, 7 and 13 was carried out, and clinical data, oxygen requirements, intensive care unit (ICU) admission, steroid treatment, and laboratory data were collected for each time point; patients were classified in severity classes on the basis of oxygen requirement, ICU admission, limitation of therapeutic effort and mortality (S1 Table), as in [9].

### Flow cytometric peripheral blood lymphocyte analysis

Peripheral blood samples were collected in EDTA-containing tubes (Becton Dickinson). Cell staining was performed on whole blood samples using a direct immunofluorescence method with erythrocytes lysis and washing. Cells were stained with a panel of four 8-colour antibody combinations (S2 and S3 Tables). Clone and isotypes and detailed in Supplementary Table 2. The antibodies were used at the dilution recommended by the manufacturers. Acquisition was performed using BD FACSCanto-II flow cytometer (BD Biosciences, San José, CA) and analysis done with BD FACSDiva 8 software (BD Biosciences, San José, CA). The absolute numbers of subsets were calculated by multiplying their percentage by the total lymphocyte number obtained from an ABX MICROS 60 device (HORIBA ABX SAS, Montpellier, France). BD CompBeads (BD Biosciences) were used for compensation settings. Cytometer performances were checked daily using CS&T IVD beads (BD Biosciences).

Flow cytometric monocyte HLA-DR expression analysis was performed as described in S1 File.

### Statistical analysis

Hierarchical cluster analysis with the Ward method [10] was used to identify groups of SARS-CoV-2 infected patients on the basis of immunophenotypic profiling; the age of the patients was included in the model (S1 File).

## Results

Median age of the patients was 70 years (IQR [56-78]), and most were male (n=74, 59%); median body mass unit (BMI) was 27 kg/m^2^ (IQR [23-32]. Mean time from symptom onset to hospital admission was 11 (sd 5.6) days. Overall, 68 (54.4%) patients were classified as severe, 41 (32.8%) have been admitted to ICU during their follow-up and 14 (11.2%) deceased; 51 (40.8%) patients were treated with corticosteroids during the follow-up (Table 1).

**Table 1.** Description of the population.

|  | Study population |
| --- | --- |
| n | 125 |
| Men, n (%) | 74 (59 %) |
| Women, n (%) | 51 (41 %) |
| Time from symptom onset to first biological sample : mean (sd) | 11 (5.6) |
| Age : mean (sd) | 67 (16.7) |
| Age : median [IQR] | 70 [55.6 ; 78.5] |
| BMI : mean (sd) | 27.4 (5.9) |
| BMI : median [IQR] | 27.1 [23.3 ; 31.9] |
| CRP : mean (sd) | 84 (79) |
| CRP : median [IQR] | 61 [25 ; 133] |
| Mortality, n (%) | 14 (11.2 %) |
| Severe COVID19<sup>1</sup>, n (%) | 68 (54.4 %) |
| ICU admission, n (%) | 41 (32.8 %) |
| Oxygen requirement, n (%) | 87 (69.6 %) |
| Limitation of therapeutic effort (LTE), n (%) | 3 (2.4 %) |
| Treated with corticosteroids, n (%) | 51 (40.8 %) |

IQR : interquartile range
<sup>1</sup> Severe Covid19 defined as: O<sub>2</sub><2L/min, ICU admission, LTE, decease

The lymphocyte subpopulation analysis performed on the day of hospital admission is summarized in Table 2; immunophenotype of cell subsets is detailed in S4 Table. Lymphopenia was observed, with a median value of total lymphocyte count of 0.9 G/L (IQR [0.6-1.3]), affecting CD3^+^ cells (median 0.6 G/L [0.4-0.9]), CD4^+^ cells (median 0.4 G/L [0.2-0.6]), CD8^+^ cells (median 0.2 G/L [0.1-0.3]), NK cells (median 0.1 G/L [0.1-0.2]), and CD19^+^ cells (median 0.1 G/L [0.1-0.2]). Other characteristics found in the study population were: effector CD8^+^CCR7^-^ T cells above normal value (median 64 % [49-81]), elevated levels of CD19^+^CD38^high^ plasmablasts (median 7 % [3-18]), HLA-DR molecules on CD14^+^ monocytes (mHLA-DR) in normal ranges (median 35952 AB/C [19850-50973]) [5] [11] [12].

**Table 2.**
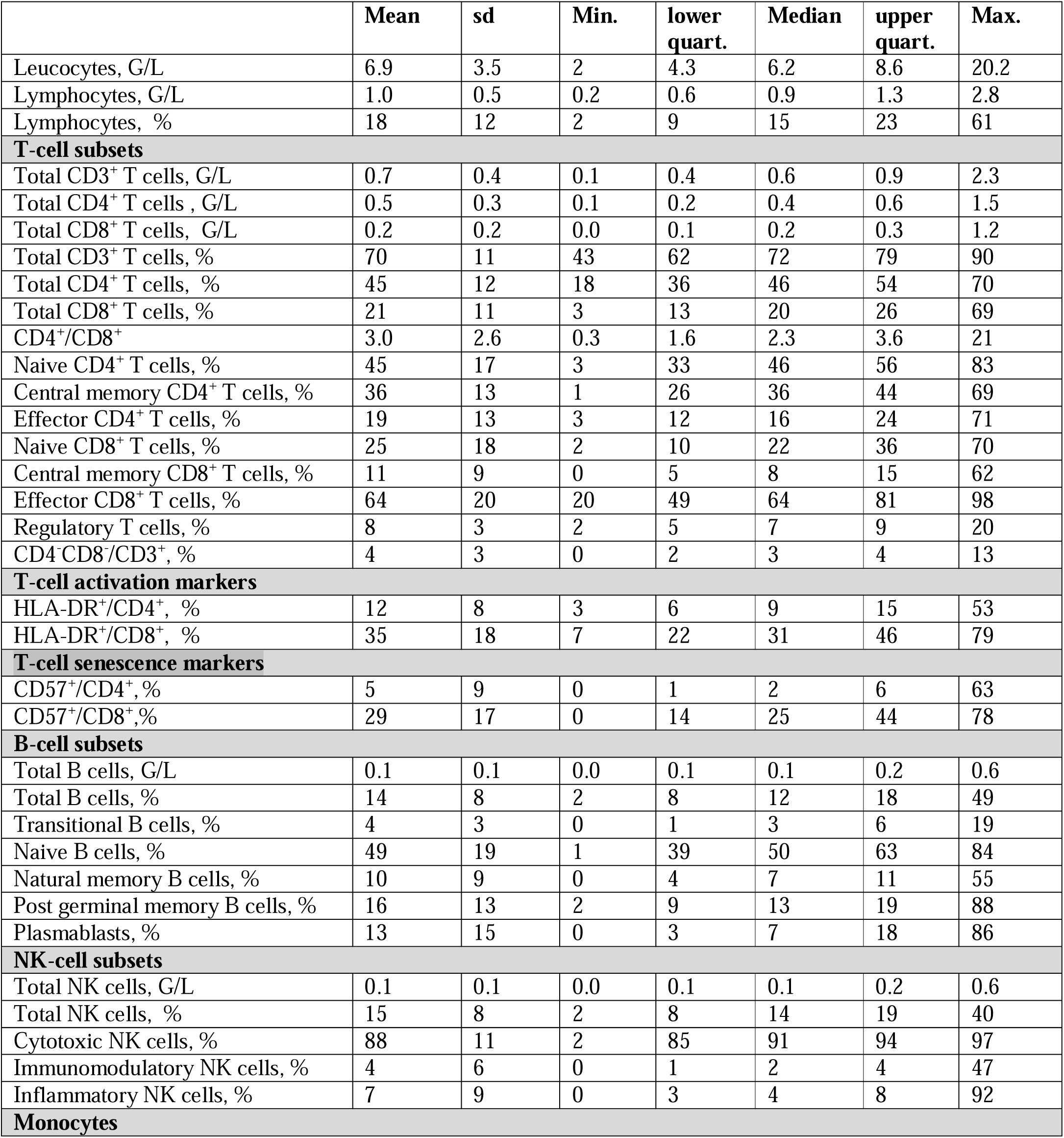
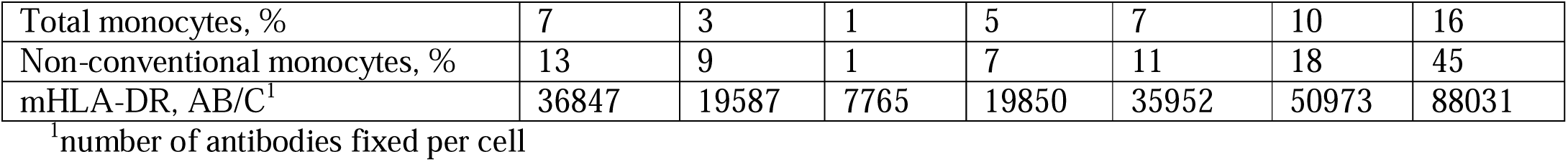
Description of cellular subpopulations.

Five clusters of patients were identified regarding immunophenotypic profile (S2 Fig). Statistical analysis of each cellular subpopulation is reported in S5 Table; the main characteristics of patients in the different clusters are described in Table 3. Some characteristics were observed in all the clusters, such as lymphopenia, an elevated level of effector CD8^+^CCR7^-^ T cells (with extremely high levels in clusters 4 and 5) and an elevated level of plasmablasts (with extremely high levels in cluster 3). Patients in cluster 1 were the youngest patients (mean age 56), with low T-cell activation (HLA-DR^+^/CD4^+^ mean 7 % and HLA-DR^+^/CD8^+^ mean 21 %) and very low T-cell senescence (CD57^+^/CD4^+^ mean 2 % and CD57^+^/CD8^+^ mean 14 %); no mortality was observed in this cluster. Patients of cluster 2 exhibited high T-cell activation (HLA-DR^+^/CD4^+^ mean 11 % and HLA-DR^+^/CD8^+^ mean 34 %) and high level of senescent T CD8^+^ cells (CD57^+^/CD8^+^ mean 33 %). Cluster 3 was specifically characterized by an extremely elevated level of CD19^+^CD38^high^ plasmablasts (mean 33 %); T-cell activation was very high (HLA-DR^+^/CD4^+^ mean 17 %; HLA-DR^+^/CD8^+^ mean 49 %), with very low level of senescent T cells (CD57^+^/CD4^+^ mean 2 % and CD57^+^/CD8^+^ mean 18 %). Cluster 4 was characterized by an extremely high level of effector CD8^+^CCR7^-^ T cells (mean 85 %), a very high level of T-cell activation (HLA-DR^+^/CD4^+^ mean 16 %; HLA-DR^+^/CD8^+^ mean 37 %) and T-cell senescence (CD57^+^/CD4^+^ mean 19 % and CD57^+^/CD8^+^ mean 52 %). Similarly to cluster 4, cluster 5 was characterized by an extremely high level of effector CD8^+^CCR7^-^ T cells (mean 84 %), a very high level of T-cell activation (HLA-DR^+^/CD4^+^ mean 14 %; HLA-DR^+^/CD8^+^ mean 48 %) and CD8^+^ T-cell senescence (CD57^+^/CD8^+^ mean 44 %); patients in cluster 5 were the oldest (mean age 79). To note, cluster 4 was the only cluster characterized by normal levels of CD8^+^ cells (mean 0.4 G/L) and a high level of effector CD4^+^CCR7^-^ T cells (mean 39 %) (Table 3, Fig 1, S6 Table).

**Table 3.**
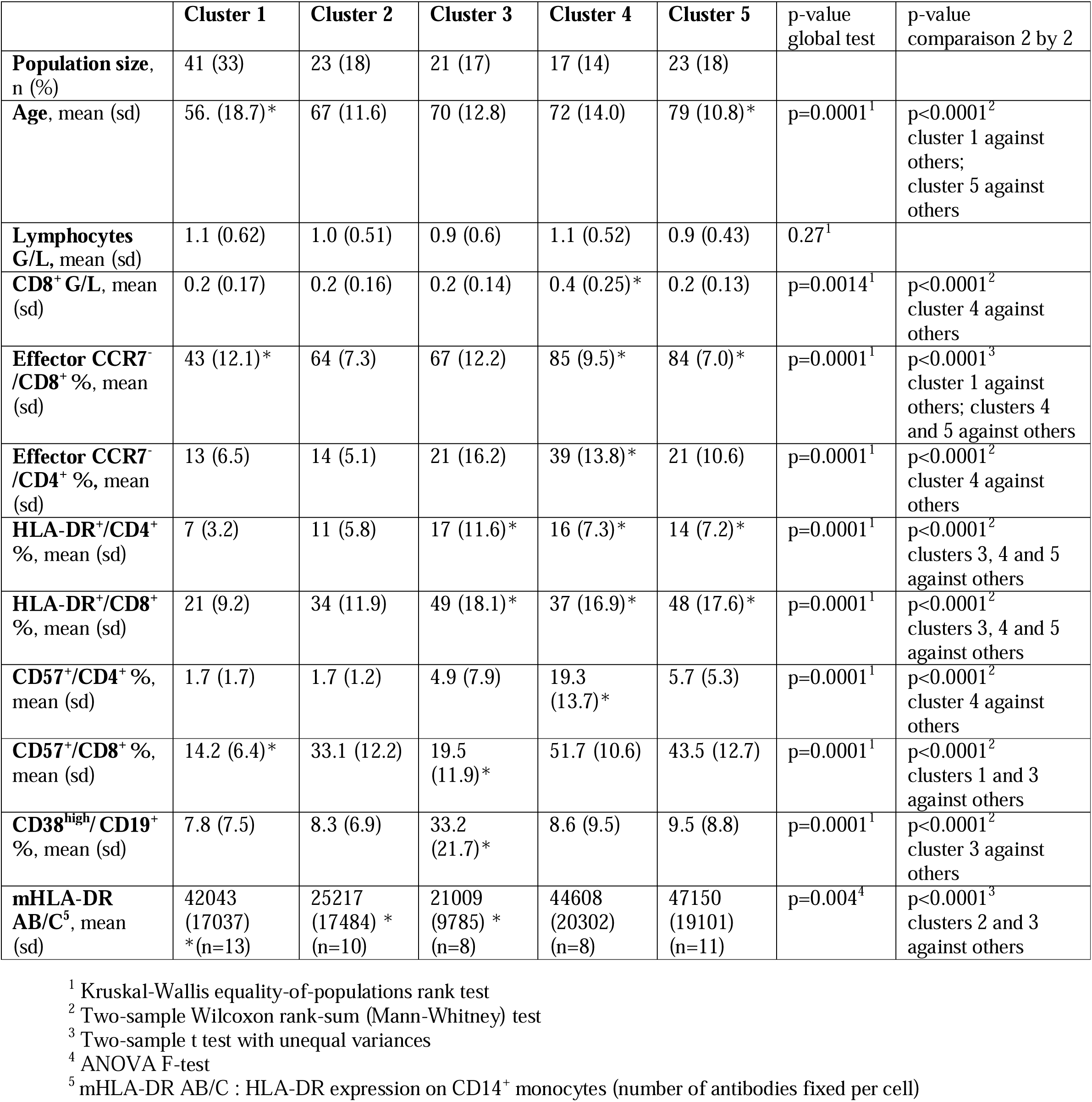
Clusters of SARS-CoV-2 infected patients according to immunophenotypic profiling.

**Fig 1.**
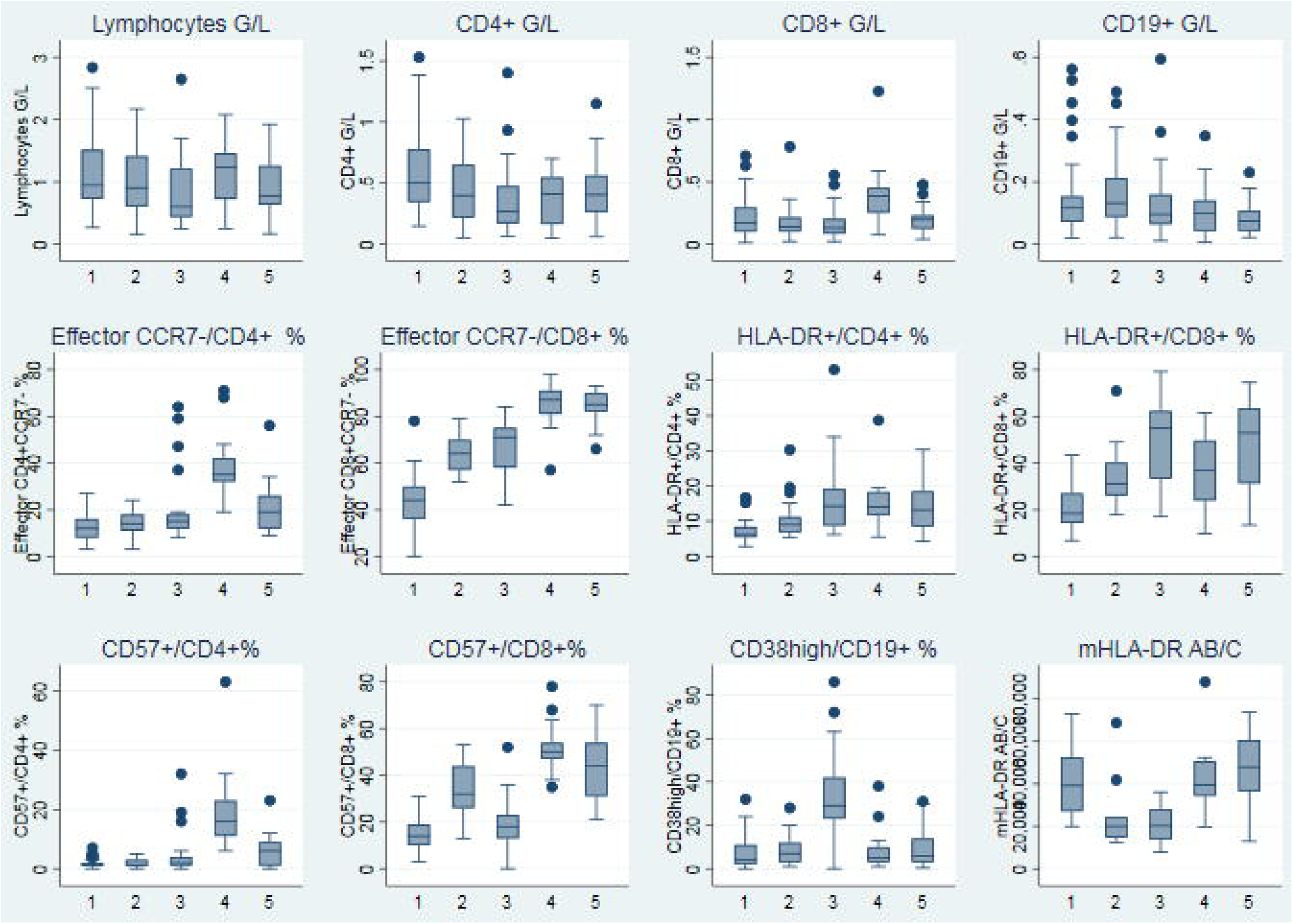
Boxplots representing the major characteristics of the five clusters of patients.

Concerning patient characteristics, there was no statistically significant difference among the clusters for sex, BMI, C-Reactive Protein (CRP), classes of severity, O_2_ requirement and ICU admission. However, a significant difference was observed concerning mortality, with a higher death rate in clusters 2 and 5 compared with the other clusters (Table 4).

**Table 4.** Overall comparison of clinical and biological characteristics between the clusters.

|  | Cluster 1 | Cluster 2 | Cluster 3 | Cluster 4 | Cluster 5 | p-value |
| --- | --- | --- | --- | --- | --- | --- |
| Men, n (%) | 24 (58.5 %) | 13 (56.5 %) | 14 (66.7 %) | 12 (70.6 %) | 11 (47.8 %) | 0.630<sup>1</sup> |
| BMI, kg/m<sup>2</sup>, mean (sd) (n=94 ) | 26.5 (6.3) | 28.4 (4.7) | 28.6 (5.8) | 27.5 (7.2) | 27.2 (6.2) | 0.745<sup>2</sup> |
| CRP, mg/L, median [IQR] (n=99 ) | 41[18;100] | 95[31;155] | 84[18;131] | 55.5[19.5;121] | 62.5[48;151.5] | 0.641<sup>3</sup> |
| Mortality, dead, n (%) | 0 (0 %) | 4 (17.4 %) | 2 (9.5 %) | 2 (11.8 %) | 6 (26.1 %) | 0.005<sup>1</sup> cluster 1 against others ;<br>0.006<sup>1</sup> clusters 2 and 5 against others |
| Severe Covid19<sup>4</sup>, n (%) | 18 (43.9 %) | 14 (60.9 %) | 16 (76.2 %) | 9 (52.9 %) | 11 (47.8 %) | 0.154<sup>1</sup> |
| ICU admission, n (%) | 11 (26.8 %) | 8 (34.8 %) | 12 (57.1 %) | 5 (29.4 %) | 5 (21.7 %) | 0.121<sup>1</sup> |
| Oxygen requirement, n (%) | 27 (65.8 %) | 17 (73.9 %) | 16 (76.2 %) | 14 (82.3 %) | 13 (56.5 %) | 0.426<sup>1</sup> |

<sup>1</sup> Fisher exact test
<sup>2</sup> F-test Anova due to normality distribution
<sup>3</sup> Kruskal Wallis test due to non-normality distribution
<sup>4</sup> Severe Covid19 defined as: O<sub>2</sub><2L/min, ICU admission, LTE, decease

## Discussion

This study had the objective of performing a fine analysis of lymphocyte subsets in SARS-CoV-2 infected patients hospitalized in Grenoble Alpes University Hospital between March and September 2020. Our results strengthen previous studies showing heterogeneous profiles of SARS-CoV-2 infected patients obtained by unsupervised clustering and pointing out that disease severity may be associated with different profiles of immune response [6] [7] [8]. Our study focuses on lymphocyte subpopulations, in patients recruited the day of their admission to the hospital and not treated yet. Interestingly, phenotypic clusters of immune response did not exhibit statistically significant differences neither concerning sex, BMI and CRP (parameters that are usually associated with a more severe outcome), nor with high-flow oxygen requirement and ICU admission; however, different mortality outcome could be pointed out.

Even if some characteristics were similar among the different clusters, such as lymphopenia, and an elevated level of effector CD8^+^CCR7^-^ T cells, some other characteristics were very different : low lymphocyte activation and senescence in cluster 1, extremely elevated level of plasmablasts in cluster 3, high CD4^+^ and CD8^+^ T-cell activation in clusters 3, 4 and 5, and high CD8^+^ T-cell senescence in clusters 2, 4 and 5. Of note, the expression of HLA-DR molecules on circulating monocytes was in normal range in the study population, indicating that patients were mainly in a status of immunocompetence. Immunosuppression status which has been described in some studies [5] has been observed mainly in patients hospitalized in ICU. Overall, exhibiting low levels of T-cell activation seems to be associated to a better disease outcome, as described in [6]; on the other hand, exhibiting profound CD8^+^ T-cell lymphopenia, a high level of CD4^+^ and CD8^+^ T-cell activation and a high level of CD8^+^ T-cell senescence seems to be globally associated with a higher mortality outcome.

Patients of our study were all hospitalized and recruited between March and September 2020. Therefore, we described here the immunophenotypic subset profiles of only severe cases from the first wave. It would be interesting to explore the profiles of not hospitalized patients with mild pathology, and those of patients infected with new circulating SARS-CoV-2 variants.

Our study suggests that some lymphocyte parameters might be useful to physicians to better characterize patients at hospital admission; in particular, identification of patients with potential mild (with low levels of T-cell activation) or very serious (with profound CD8^+^ T-cell lymphopenia, a high level of CD4^+^ and CD8^+^ T-cell activation and a high level of CD8^+^ T-cell senescence) evolution of the pathology could be helpful in order to treat them earlier and more appropriately. In this perspective, specific studies evaluating T-cell activation and senescence in a longitudinal patient follow-up are certainly needed.

## Data Availability

All data are available in the manuscript

## Acknowledgments

The authors would like to thank Ghislaine Del Vecchio, Richard Di Schiena, Laure Dusset, Claire Gasquez, Frédérique Martinez, Karine Nicolino and Véronique Bergerot who provided technical support and Sylvie Berthier who helped to collect clinical data.

## Supporting information

**S1 File. Supporting methods**.

**S1 Fig. Enrollment and inclusion of the patients. S2 Fig. Clustering of the patients**.

**S1 Table. Classification of severity**.

**S2 Table. Panel of four antibody combinations used in the study. S3 Table. Characteristics of antibodies**.

**S4 Table. Cell subsets and corresponding immunophenotypes**.

**S5 Table. Overall comparison of cellular subpopulations between the clusters. S6 Table. Major characteristics of the five clusters of patients**.

## Notes

### Competing Interest Statement

The authors have declared no competing interest.

### Clinical Trial

NCT04596098

### Funding Statement

No external funding was received for this work

### Author Declarations

Ethical approval for the prospective study was given by the relevant ethics committee (Comite de Protection des Personnes Ile de France II, IDRCB: 2020-A00904-35) CPP Ile de France II 60 Boulevard du Marechal Martial Valin, CS 21623 75509 PARIS CEDEX 15 Bureau : President : Stephane DONNADIEU Vice-Presidente : Marie-France MAMZER-BRUNEEL Secretaires : Laura CHEVREAU, Pierre COLONNA Membres : C. ARDIOT, C. BALLOUARD, J.-L. BRESSON, C. BROISSAND, M. DE FALLOIS, A.GOUEL, L.GUEST, M-C LAI, M.LARGEAU, C. MARTINEAUX, M.SEHAN, L.VALENTINO, M. WACK, R.JACOB-VESTLING Secretariat : Nora VESTRIS

