## Supplementary Methods, Figures and Tables for "Fine analysis of lymphocyte subpopulations in SARS-CoV-2 infected patients: toward a differential profiling of patients with severe outcome"

### Flow cytometric monocyte HLA-DR expression analysis

Peripheral blood samples were collected in EDTA-containing tubes which were kept on ice and rapidly routed to the laboratory. Whole blood (50 µl) was stained with 20 µl of QuantiBrite anti-HLA-DR/Monocyte mixture (QuantiBrite anti-HLA-DR PE (clone L243)/Anti-monocytes (CD14) PerCP-Cy5.5 (clone MΦP9), Becton Dickinson, San José, CA) at room temperature for 30 min in the dark. Samples were the lysed using the FACS Lysing solution (Becton Dickinson) for 15 min. After a washing step, cells were analyzed with BD FACSCanto-II flow cytometer and FACSDiva software version 8 (BD Biosciences, San José, CA). Monocytes were first gated out from other cells on the basis of CD14 expression and mHLA-DR expression was then measured on their surface (mono-parametric histogram) as median of fluorescence intensity related to the entire monocyte population (as recommended by manufacturer). These results were then transformed in AB/C (number of antibodies fixed per cell) thanks to calibrated PE-beads (BD QuantiBrite-PE Beads, Becton Dickinson).

### Statistical analysis

Primary objective was analyzed using a non-supervised analysis: hierarchical ascendant clustering (HAC) [13]. The goal of this method is to cluster the entire analyzed population. This use an iterative way to group observations in clusters, by a distance and an aggregation method. The end of iterations results in a unique cluster made up of the whole observations. We can illustrate results with a dendrogram.

HAC need two parameters a priori:

- A distance: in this study the Euclidian distance

- An aggregation method: in this study the Ward's linkage method [10]

HAC results in all the possible clusters with the previously parameters, and defined variables. In our case, variables are the following: Leucocytes G/L, Lymphocytes G/L, total $CD3^+$ T cells G/L, total $CD4^+$ T cells G/L, total $CD8^+$ T cells G/L, total $CD3^+$ T cells %, total $CD4^+$ T cells %, total $CD8^+$ T cells %, $CD4^+/CD8^+$, CD3-CD56+ %, naive $CD4^+$ T cells %, central memory $CD4^+$ T cells %, effector $CD4^+$ T cells %, naive $CD8^+$ T cells %, central memory $CD8^+$ T cells %, effector $CD8^+$ T cells %, CD8 $CDRA^+$ CCR7 %, regulatory T cells %, $CD4^-CD8^-/CD3^+$ %, $CD57^+/CD4^+$ %, $CD57^+/CD8^+$ %, CD57 %, $CD56^+/CD4^+$ %, $CD56^+/CD8^+$ %, HLA-$DR^+/CD4^+$ %, HLA-$DR^+/CD8^+$ %, $CD25^+/CD4^+$ %, $CD25^+/CD8^+$ %, total B cells G/L total B cells %, transitional B cells %, naive B cells %, natural memory B cells %, post germinal memory B cells %, plasmablasts %, post germinal switched memory B cells %, total NK cells G/L, total NK cells %, cytotoxic NK cells %, inflammatory NK cells %, immunomodulatory NK cells %, $CD16^-$ $CD56^-$ %, total monocytes %, non-conventional monocytes %. We added age, which might be a variation factor for some of the lymphocyte subpopulations [12].

HAC was implemented using Stata 15 (StataCorp, College Station, TX, USA), with the command cluster 'wardslinkage'. The optimal number of clusters was choosing using selection criterions of Calinski-Harabasz and Duda-Hart, with the following command: 'cluster stop'.

A biological meaning of the clusters was analyzed by screening the values of every parameter, between all clusters. ANOVA F-test was conducted for parameters with a Gaussian distribution, and a non-parametric Mann-Whitney test was run for other distributions. For significant results, a specific cluster by cluster tests were conducted to identify the significantly different cluster. This was conducted using Student tests for

Gaussian distributions, Wilcoxson tests for others. Only high significant results were discussed, to take part of the risk of error inflation, due to multiplicity of tests.

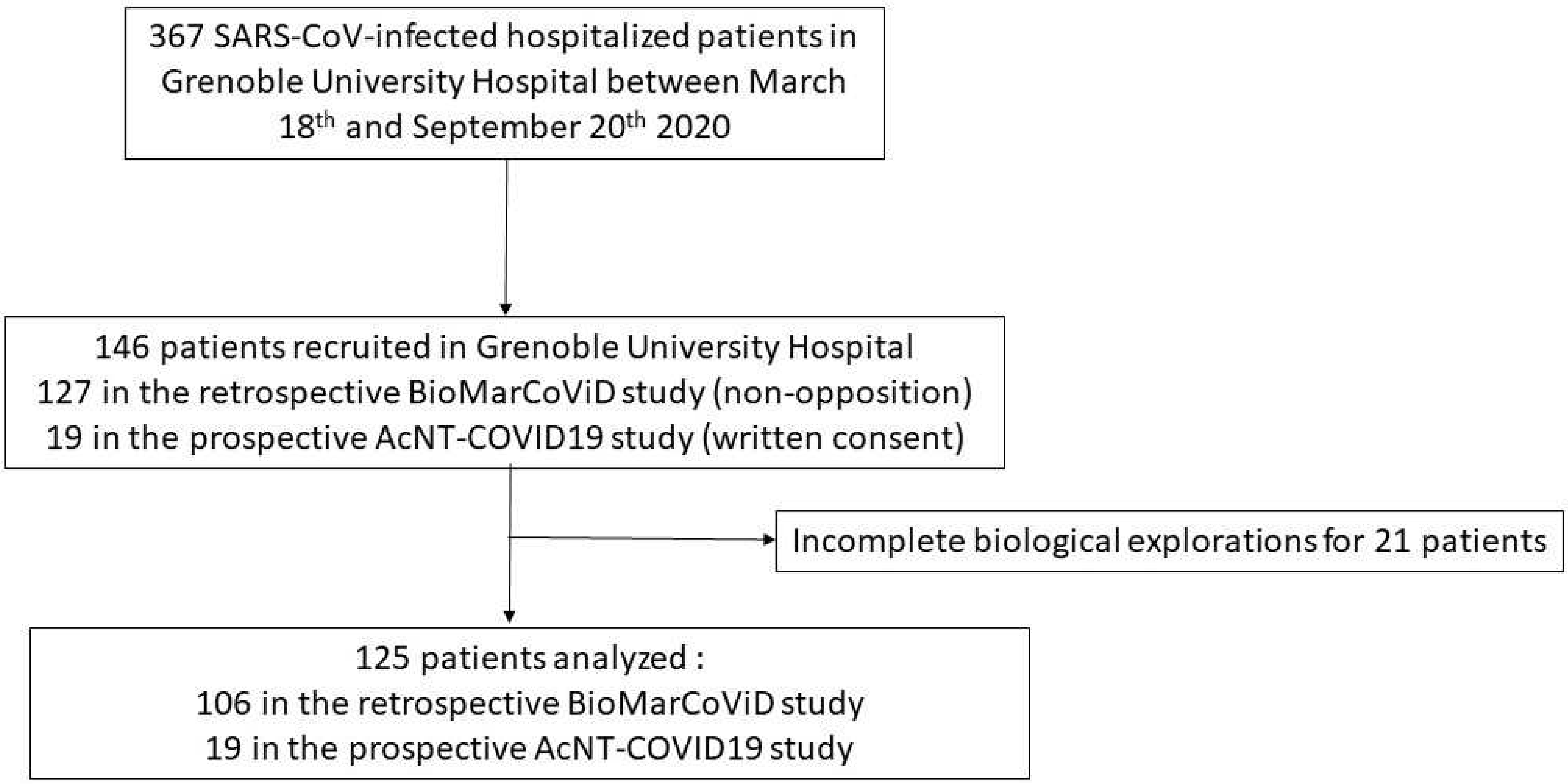
367 SARS-CoV-infected hospitalized patients in Grenoble University Hospital between March 18th and September 20th 2020
146 patients recruited in Grenoble University Hospital
127 in the retrospective BioMarCoViD study (non-opposition)
19 in the prospective AcNT-COVID19 study (written consent)
Incomplete biological explorations for 21 patients
125 patients analyzed :
106 in the retrospective BioMarCoViD study
19 in the prospective AcNT-COVID19 study

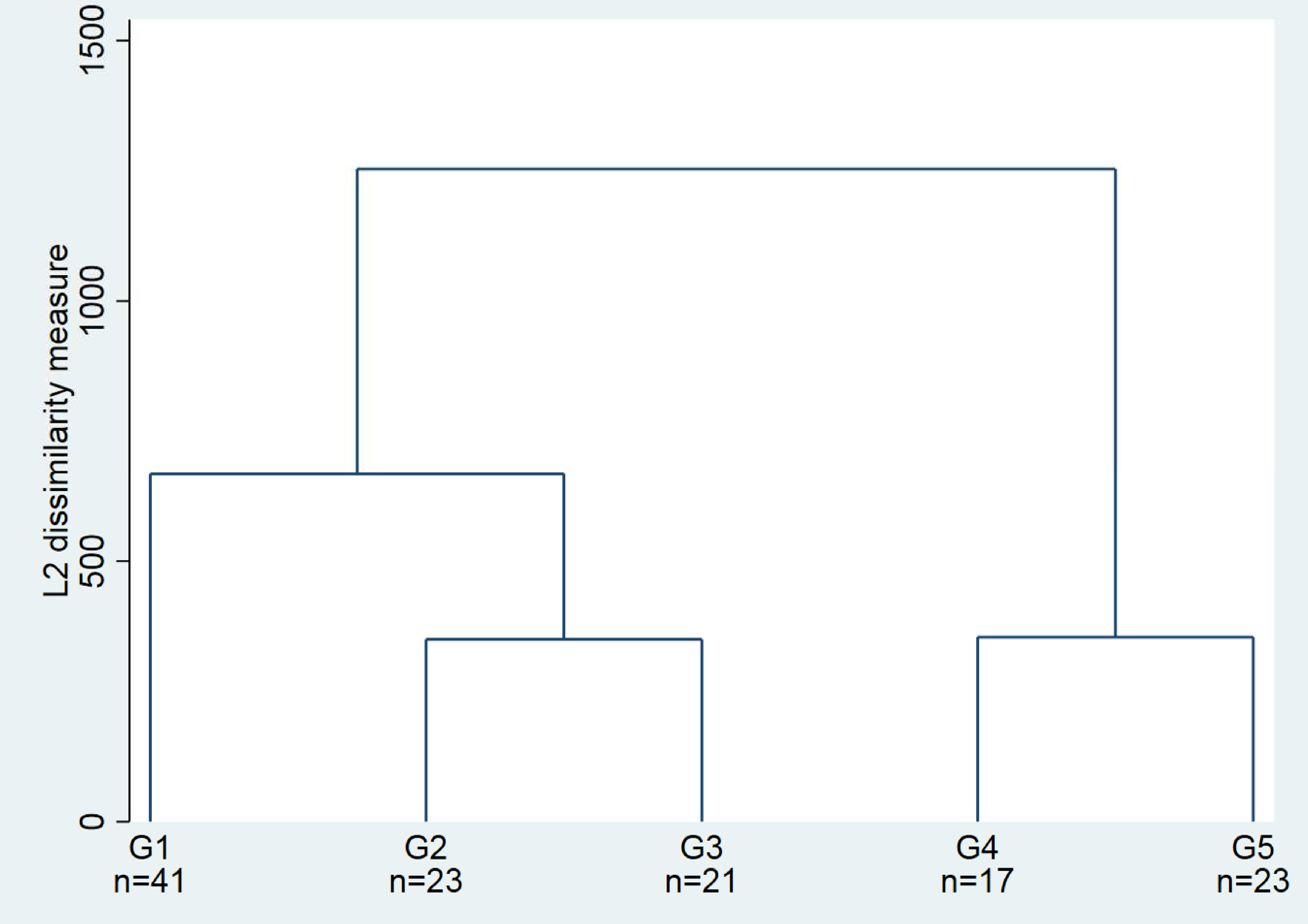
L2 dissimilarity measure
1500
1000
500
0
G1
n=41
G2
n=23
G3
n=21
G4
n=17
G5
n=23

**S1 Table.** Classification of severity.

| **Characteristics** | **Class of severity** |
|---|---|
| no $O_2$ requirement | mild disease |
| $O_2 \leq$ 2L/min | mild disease |
| $O_2 >$ 2L/min | severe disease |
| ICU admission | severe disease |
| LTE | severe disease |
| decease | severe disease |

LTE : Limitation of Therapeutic Effort

**S2 Table.** Panel of four antibody combinations used in the study.

| Fluoro-chrome / Tube | FITC | PE | PerCP or PerCP-Cy5.5 | PE-Cy7 | APC | APC-Cy7 or APC-H7 | V450 | V500 |
|---|---|---|---|---|---|---|---|---|
| 1 | CD3 | CD56 and CD16 | CD45 | CD4 | CD19 | CD8 | HLA-DR | x |
| 2 | CD57 | CD8 | CD45 | CD3 | CD45RA | x | CCR7 | x |
| 3 | IgD | CD10 | CD38 | CD27 | IgM | CD19 | x | x |
| 4 | CD3 | CD127 | CD45 | CD56 | CD25 | CD16 | CD7 | CD45 |

**S3 Table.** Characteristics of antibodies.

| CD | Fluorochrome | Clone | Isotype | Supplier | Catalogue N° |
|---|---|---|---|---|---|
| CD3 | FITC | SK7 | Ms IgG1, κ | BD Biosciences | 644611 |
| CD4 | PE-Cy7 | SK3 | Ms IgG1, κ | BD Biosciences | 644611 |
| CD8 | APC-H7 | SK1 | Ms IgG1, κ | BD Biosciences | 644611 |
| CD19 | APC | SJ25C1 | Ms IgG1, κ | BD Biosciences | 644611 |
| CD16 | PE | B73.1 | Ms IgG1, κ | BD Biosciences | 644611 |
| CD56 | PE | NCAM16.2 | Ms IgG2b, κ | BD Biosciences | 644611 |
| CD45 | PerCP | 2D1 | Ms IgG1, κ | BD Biosciences | 644611 |
| HLA-DR | V450 | L243 | Ms IgG2a, κ | BD Biosciences | 655874 |
| CD8 | PE | B9.11 | Ms IgG1 | Beckman Coulter | A07757 |
| CD4 | PerCP-Cy5.5 | SK3 | Ms IgG1, κ | BD Biosciences | 332772 |
| CD3 | PE-Cy7 | UCHT1 | Ms IgG1 | Beckman Coulter | 737657 |
| CD45RA | APC | HI100 | Ms IgG2b, κ | BD Biosciences | 550855 |
| CD197 | VB421 | 150503 | Ms IgG2a | BD Biosciences | 562555 |
| IgD | FITC | polyclonal | rabit anti-human | Agilent technologies | F018901 |
| CD10 | PE | HI10a | Ms IgG1, κ | BD Biosciences | 332776 |
| CD38 | PerCP-Cy5.5 | HIT2 | Ms IgG1, κ | BD Biosciences | 551400 |
| CD27 | PE-Cy7 | 1A4CD27 | Ms IgG1 | Beckman Coulter | B49205 |
| IgM | APC | G20-127 | Ms IgG1, κ | BD Biosciences | 551062 |
| CD19 | APC-H7 | SJ25C1 | Ms IgG1, κ | BD Biosciences | 641395 |
| CD127 | PE | HIL-7R-M21 | Ms IgG1, κ | BD Biosciences | 557938 |
| CD56 | PE-Cy7 | N901 (NKH-1) | Ms IgG1 | BD Biosciences | A21692 |
| CD25 | APC | 2A3 | Ms IgG1, κ | BD Biosciences | 340907 |
| CD16 | APC-H7 | 3G8 | Ms IgG1, κ | BD Biosciences | 560195 |
| CD7 | V450 | T701 | Ms IgG1, κ | BD Biosciences | 642916 |
| CD45 | V500 | HI30 | Ms IgG1, κ | BD Biosciences | 560777 |

**S4 Table.** Cell subsets and corresponding immunophenotypes.

| **Cell subset** | **Immunophenotype** |
|---|---|
| **T-cell subsets** | |
| Total $CD4^+$ T cells | $CD3^+$ $CD4^+$ |
| Naive $CD4^+$ T cells | $CD45RA^+$ $CCR7^-$ |
| Central memory $CD4^+$ T cells | $CD45RA^-$ $CCR7^-$ |
| Effector $CD4^+$ T cells | $CD45RA^+$ $CCR7^-$ |
| Regulatory T cells | $CD4^+$ $CD127^{low}$ $CD25^{high}$ |
| Total $CD8^+$ T cells | $CD3^+$ $CD8^+$ |
| Naive $CD8^+$ T cells | $CD45RA^+$ $CCR7^-$ |
| Central memory $CD8^+$ T cells | $CD45RA^-$ $CCR7^-$ |
| Effector $CD8^+$ T cells | $CD45RA^{+/-}$ $CCR7^-$ |
| **B-cell subsets** | |
| Total B cells | $CD19^+$ |
| Transitional B cells | $IgD^+$ $CD27^-$ $CD10^+$ $CD38^{high}$ |
| Naive B cells | $IgD^+$ $CD27^-$ $CD10^-$ $CD38^{low}$ |
| Natural memory B cells | $IgD^+$ $CD27^+$ |
| Post germinal memory B cells | $IgD^-$ $CD27^+$ $CD38^{low}$ |
| Plasmablasts | $IgD^-$ $CD27^{high}$ $CD38^{high}$ |
| **NK cells** | |
| Total NK cells | $CD56^+$ or $CD16^+$ and $CD3^-$ |
| Cytotoxic NK cells | $CD56^+$ $CD16^+$ $CD3^-$ |
| Immunomodulatory NK cells | $CD56^-$ $CD16^+$ $CD3^-$ |
| Inflammatory NK cells | $CD56^+$ $CD16^-$ $CD3^-$ |
| **Monocytes** | |
| Total monocytes | $CD45^{high}$ $SSC^{intermediate}$ |
| Non-conventional monocytes | $CD16^+$ |

**S5 Table.** Overall comparison of cellular subpopulations between the clusters.

| | Cluster 1 | Cluster 2 | Cluster 3 | Cluster 4 | Cluster 5 | p-value |
|---|---|---|---|---|---|---|
| Leucocytes, G/L | 5.7 [3.7;8.2] | 6.3 [4.6;10.3] | 8.5 [5.9;10.8] | 5.4 [4.1;7.8] | 6.1 [3.9;7.1] | 0.089[1] |
| Lymphocytes, G/L | 0.9 [0.7;1.5] | 0.9 [0.6;1.4] | 0.6 [0.4;1.2] | 1.2 [0.7;1.5] | 0.8 [0.6;1.3] | 0.27[1] |
| Lymphocytes, % | 18 [12;28] | 1 4 [7;23] | 7 [6;15] | 19 [11;24] | 17 [10;23] | 0.018[1] |
| **T-cell subsets** | | | | | | |
| Total $CD3^+$ T cells, G/L | 0.7 [0.5;1.1] | 0.6 [0.4;0.9] | 0.4 [0.3;0.7] | 0.8 [0.6;1] | 0.6 [0.4;0.9] | 0.11[1] |
| Total $CD4^+$ T cells , G/L | 0.5 [0.3;0.8] | 0.4 [0.2;0.6] | 0.3 [0.2;0.5] | 0.4 [0.2;0.5] | 0.4 [0.3;0.6] | 0.049[1] |
| Total $CD8^+$ T cells, G/L | 0.2 [0.1;0.3] | 0.1 [0.1;0.2] | 0.1 [0.1;0.2] | 0.4 [0.2;0.5] | 0.2 [0.1;0.2] | 0.001[1] |
| Total $CD3^+$ T cells, % | 72 [67;81] | 63 [55;71] | 67 [59;74] | 77 [74;82] | 74 [66;83] | p<0.001[1] |
| Total $CD4^+$ T cells, % | 52 (10) | 43 (10) | 43 (12) | 32 (9) | 48 (10) | p<0.001[2] |
| Total $CD8^+$ T cells, % | 17 (8) | 17 (7) | 19 (7) | 38 (14) | 22 (8) | p<0.001[2] |
| $CD4^+/CD8^+$ | 3.1[2;4.9] | 2.8 [1.7;3.5] | 2 [1.5;3.4] | 0.9 [0.8;1.5] | 1.9 [1.6;3.6] | p<0.001[1] |
| Naive $CD4^+$ T cells, % | 53 (16) | 36 (12) | 47 (18) | 25 (10) | 51 (13) | p<0.001[2] |
| Central memory $CD4^+$ T cells, % | 34 (12) | 49 (11) | 32 (8) | 36 (11) | 28 (11) | p<0.001[2] |
| Effector $CD4^+$ T cells, % | 12 [8;16] | 14 [11;18] | 15 [12;18] | 35 [32;42] | 19 [12;26] | p<0.001[1] |
| Naive $CD8^+$ T cells, % | 46 [32;55] | 21 [15;27] | 22 [15;33] | 7 [5;11] | 8 [6;14] | p<0.001[1] |
| Central memory $CD8^+$ T cells, % | 11 [7;19] | 15 [10;18] | 6 [5;12] | 5 [3;8] | 4 [3;7] | p<0.001[1] |
| Effector $CD8^+$ T cells, % | 44 [36;50] | 64 [57;70] | 71 [58;75] | 87 [81;91] | 85 [82;90] | p<0.001[1] |
| Regulatory T cells, % | 7 [5;8] | 9 [7;10] | 8 [6;10] | 7 [5;9] | 7 [6;8] | 0.049[1] |
| $CD4^-CD8^-/CD3^+$, % | 2 [2;4] | 3 [2;5] | 2 [2;3] | 4 [2;4] | 2 [1;6] | 0.463[1] |
| **T-cell activation markers** | | | | | | |
| $HLA\text{-}DR^+/CD4^+$, % | 6 [5;8] | 9 [7;11] | 14 [9;19] | 14 [12;18] | 13 [8;18] | p<0.001[1] |
| $HLA\text{-}DR^+/CD8^+$, % | 18 [14;27] | 31 [26;40] | 55 [33;62] | 37 [24;50] | 53 [31;63] | p<0.001[1] |
| **T-cell senescence markers** | | | | | | |
| $CD57^+/CD4^+$, % | 1 [1;2] | 1 [1;3] | 2 [1;4] | 16 [11;23] | 6 [1;9] | p<0.001[1] |
| $CD57^+/CD8^+$, % | 14 [10;19] | 32 [26;44] | 18 [13;23] | 50 [47;54] | 44 [31;54] | p<0.001[1] |
| **B-cell subsets** | | | | | | |
| Total B cells, G/L | 0.1 [0.1;0.2] | 0.1 [0.1;0.2] | 0.1 [0.1;0.2] | 0.1 [0.0;0.1] | 0.1 [0.0;0.1] | 0.049[1] |
| Total B cells, % | 12 [8;17] | 17 [12;23] | 17 [10;24] | 10 [7;11] | 9 [6;12] | 0.001[1] |
| Transitional B cells, % | 4 [2;6] | 4 [2;6] | 1 [1;4] | 5 [2;6] | 1 [1;3] | p<0.001[1] |
| Naive B cells, % | 58 (16) | 55 (14) | 31 (17) | 53 (12) | 43 (22) | p<0.001[2] |
| Natural memory B cells, % | 8 [4;12] | 8 [4;10] | 6 [3;21] | 9 [5;14] | 6 [4;9] | 0.794[1] |
| Post germinal memory B cells, % | 12 [8;18] | 14 [9;18] | 9 [7;16] | 13 [9;19] | 15 [11;35] | 0.056[1] |
| Plasmablasts, % | 4 [2;11] | 7 [3;12] | 29 [23;42] | 5 [3;10] | 6 [3;14] | p<0.001[1] |
| **NK-cell subsets** | | | | | | |
| Total NK cells, G/L | 0.1 [0.1;0.2] | 0.1 [0.1;0.2] | 0.1 [0.0;0.1] | 0.1 [0.1;0.2] | 0.1 [0.1;0.2] | 0.57[1] |
| Total NK cells, % | 12 [7;18] | 15 [9;25] | 14 [8;21] | 12 [10;18] | 14 [6;21] | 0.595[1] |
| Cytotoxic NK cells, % | 89 [85;94] | 89 [79;92] | 90 [88;94] | 92 [85;96] | 92 [90;94] | 0.105[1] |
| Inflammatory NK cells, % | 2 [1;4] | 3 [1;7] | 2 [1;4] | 2 [1;3] | 2 [2;5] | 0.661[1] |
| Immunomodulatory NK cells, % | 5 [4;9] | 6 [4;13] | 4 [3;8] | 4 [3;7] | 4 [2;6] | 0.206[1] |
| **Monocytes** | | | | | | |
| Total monocytes, % | 8.0 (3.6) | 7.5 (3.3) | 7.1 (2.7) | 7.0 (3.0) | 7.2 (3.3) | 0.779[2] |
| Non-conventional monocytes, % | 12 [7;19] | 11 [7;14] | 5 [3;12] | 13 [10;17] | 14 [10;26] | 0.003[1] |
| mHLA-DR, AB/C[3] | 42044 (17038) | 25218 (17485) | 21009 (9786) | 44608 (20302) | 47150 (19101) | 0.004[2] |
| Other | | | | | | |
| Age | 56 [44.6;72.1] | 70 [56.6;75.6] | 71 [62.3;80.6] | 72 [64.1;82.5] | 79 [70.8;89.3] | p<0.001[1] |

[1] p-value from Kruskal Wallis test due to non-normality distribution; median [IQR]

[2] p-value from F-test Anova due to normality distribution; mean (SD)

[3] number of antibodies fixed per cell

**S6 Table.** Major characteristics of the five clusters of patients.

| | Cluster 1 | Cluster 2 | Cluster 3 | Cluster 4 | Cluster 5 |
|---|---|---|---|---|---|
| **Age (mean)[1]** | 56.4 | 66.6 | 70.1 | 72.5 | 79 |
| **Lymphocytes G/L** | low | low | low | low | low |
| **$CD8^+$ T cells** | low | very low | very low | normal | low |
| **Effector $CD8^+$ T cells** | high | very high | very high | extremely high | extremely high |
| **Effector $CD4^+$ T cells** | normal | normal | normal | high | normal |
| **$CD4^+$ and CD8+ activation** | low activation | high | very high | very high | very high |
| **$CD8^+$ senescence** | low senescence | high | low senescence | very high | very high |
| **$CD4^+$ senescence** | low senescence | low senescence | low senescence | very high | high |
| **Plasmablasts** | normal/high | high | extremely high | high | high |
| **mHLA-DR** | normal | normal/low | normal/low | normal | normal |
| **Mortality[2]** | 0 | 17.4 | 9.5 | 11.1 | 26.1 |

[1] $p<0.0001$ cluster 1 against others; cluster 5 against others
[2] $p=0.005$ cluster 1 against others; $p= 0.006$ clusters 2 and 5 against others
